# Barriers and facilitators to implementing DREAMS:START intervention for sleep disturbance in dementia within the NHS: A pre-implementation study

**DOI:** 10.1101/2025.10.29.25339056

**Authors:** Vrushti Mehta, Penny Rapaport, Gill Livingston

**Author notes:** Corresponding author: Vrushti Mehta.

## Abstract

**Background:** Sleep disturbances are common in people living with dementia and negatively affect them and their family carers. Non-pharmacological interventions like DREAMS:START (Dementia Related Manual for Sleep: Strategies for Relatives) offer potential benefits but are not yet implemented widely in NHS practice.

**Aims:** To explore the barriers and facilitators to implementing DREAMS:START in UK NHS dementia services, from the perspective of healthcare professionals and intervention facilitators, using qualitative interviews.

**Method:** We interviewed 19 NHS staff - (n=10) staff with no prior DREAMS:START experience and (n=9) DREAMS:START facilitators using semi-structured interviews and analysed data using reflexive thematic analysis, organized according to the Consolidated Framework for Implementation Research (CFIR). Ethical approval was granted by the London-Camden and Kings Cross Research Ethics Committee (Reference: 20/LO/0894).

**Results:** Five key themes emerged: 1) perceived need for interventions targeting sleep in dementia; 2) importance of evidence and cost-effectiveness; 3) influence of service manager attitudes and support; 4) inter-professional interconnectedness and training; and 5) staff capacity and attitudes toward change. Participants highlighted the need for flexible, personalised delivery, robust evidence, training, supervision, and local champions to facilitate adoption.

**Conclusions:** Implementation of DREAMS:START in NHS services is feasible but organisational barriers need addressing to enhance staff capacity, and promote evidence-based, cost-effective practices. Findings from this study will inform strategies for scaling up the intervention to improve sleep outcomes for people with dementia and their carers.

## Introduction

Almost 1 million people in the United Kingdom and 55 million globally live with dementia (ARUK, 2022; WHO, 2020). At any one time, around 39% of people with dementia have sleep problems (Koren et al., 2023), with symptoms such as sleep fragmentation, night-time wandering, and daytime sleepiness (Kinnunen et al., 2017). Family members may be repeatedly woken and find it harder to fall asleep (Gao et al., 2019; Lee et al., 2010). Thus, carers find it difficult to care for relatives with disturbed sleep, and people with dementia experiencing sleep problems are more likely to move to care homes (Gibson et al., 2023). Sleep disturbances in dementia often have multiple causes, including dysregulation of the circadian rhythm, reduced exposure to sunlight and lack of physical activity, anxiety or depression, and pain from comorbid conditions (Leng et al., 2019; Livingston et al., 2018).

There is no definitive randomised controlled trial (RCT) evidence in people with dementia for widely prescribed drugs, like melatonin, benzodiazepine and non-benzodiazepine hypnotics, many of which have harmful side effects (Livingston et al., 2024; McCleery et al., 2020) and non-pharmacological approaches should be first line treatment (Livingston et al., 2017; NICE; 2018). DREAMS:START (Dementia Related Manual for Sleep: Strategies for Relatives) is the first multicomponent intervention with full RCT evidence of clinical and cost-effectiveness in improving sleep for people living at home with dementia and their family carers (Rapaport et al., 2024; Gonzalez et al., 2025). It aligns with the NICE guidelines (2018) as a “personalised multi-component sleep management approach – including sleep hygiene education, exposure to daylight, and exercise”. It comprises six sessions delivered, either in-person or online, by a non-clinical facilitator to a family carer who then implements changes with their relative and incorporates sleep hygiene, relaxation, education, exposure to daylight, a lightbox, tailored sleep bed and rise times via wrist worn actigraphy and increasing exercise and daytime activity (Amador et al, 2025).

Currently, DREAMS:START is not implemented in routine health care including National Health Service (NHS). Therefore, in this pre-implementation study we interviewed stakeholders delivering dementia support in the United Kingdom NHS to comprehensively understand their views regarding potential barriers and facilitators to implementing DREAMS:START in the NHS to help translation into real-life clinical settings (Rycroft-Malone et al., 2012 delineating any necessary adjustments, leading to more purposeful planning for full implementation (Walker et al., 2014).

## Methods

### Study Design

This was a qualitative study using semi-structured interviews with NHS professionals working with people with dementia, and facilitators of the DREAMS:START intervention.

### Ethics

The authors assert that all procedures contributing to this work comply with the ethical standards of the relevant national and institutional committees on human experimentation and with the Helsinki Declaration of 1975, as revised in 2013. The study received ethical approval from the London-Camden & Kings Cross Research Ethics Committee (Reference: 20/LO/0894) in September 2020 with an amendment agreed in April 2024 to conduct additional interviews. All participants gave written and audio recorded informed consent.

### Participants and Setting

We recruited mental health professionals with no prior DREAMS:START training from UK NHS secondary care dementia services, and facilitators who had been trained and delivered the intervention within the RCT (Amador et al., 2025). We purposively recruited to incorporate different geographical and organizational contexts and facilitators who had delivered in the intervention at different sites, number of times and varying job roles. We aimed to recruit mental health workers working in memory services, older adult community mental health teams, and integrated care team. Data collection continued until data saturation (Mwita, 2024). We identified potential health service professional participants who had expressed an interest through professional forums, NHS dementia clinical networks, and organisations focused on dementia. Once participation was confirmed, we sent the participant information sheet, consent form, DREAM:START content summary and clinical findings via email.

### Data Collection

We adhered to the COREQ (Consolidated Criteria for Reporting Qualitative Research) guidelines (Tong et al., 2007) (see Supplementary Material). VM interviewed NHS professionals working with people with dementia using a semi-structured interview guide to provide insights from health professionals on how they felt implementation may work in practice, outside research settings. We also integrated findings from previously conducted qualitative interviews with facilitators conducted as part of our main trial process evaluation. We collected demographic details. The topic guide was developed to explore how organisational and staff factors might affect the implementation of DREAMS:START. It encouraged participants to reflect on 1) sleep disturbances that people with dementia and their carers report, and the support provided; 2) interventions previously implemented in their organization and 3) funding, organisational and staff considerations for implementing DREAMS:START in the NHS (see Supplementary Material).

### Data analysis

We transcribed the interviews which had been audio-recorded, anonymised and checked transcripts for accuracy, and entered into NVivo 11 software.

The team met to develop themes using a reflexive thematic analysis approach (Braun et al., 2019). We began with familiarisation, by reading and re-reading transcripts to gain a comprehensive understanding of the data. The team met 12 times to discuss, develop and finalise themes. Ongoing discussions supporting reflexivity allowed us to question assumptions and deepen interpretations. We included the range of experiences and different voices, rather than emphasising frequency of opinions. We developed the initial codes inductively, through open coding. Once completed, we grouped related codes into broader conceptual themes informed by the 37-item Consolidated Framework for Implementation Research (CFIR) (Kirk et al., 2015) identifying both commonalities and variations across different settings. The CFIR is a widely used framework in implementation science, (see Figure 1) which organises implementation factors into five major domains – intervention characteristics (attributes of the intervention itself), outer setting (external factors like patient needs and policies), inner setting (organisational features like culture, leadership), characteristics of individuals (knowledge, beliefs, attitudes of people involved in implementation), and process (the steps taken to implement the intervention).

**Figure: 1.**
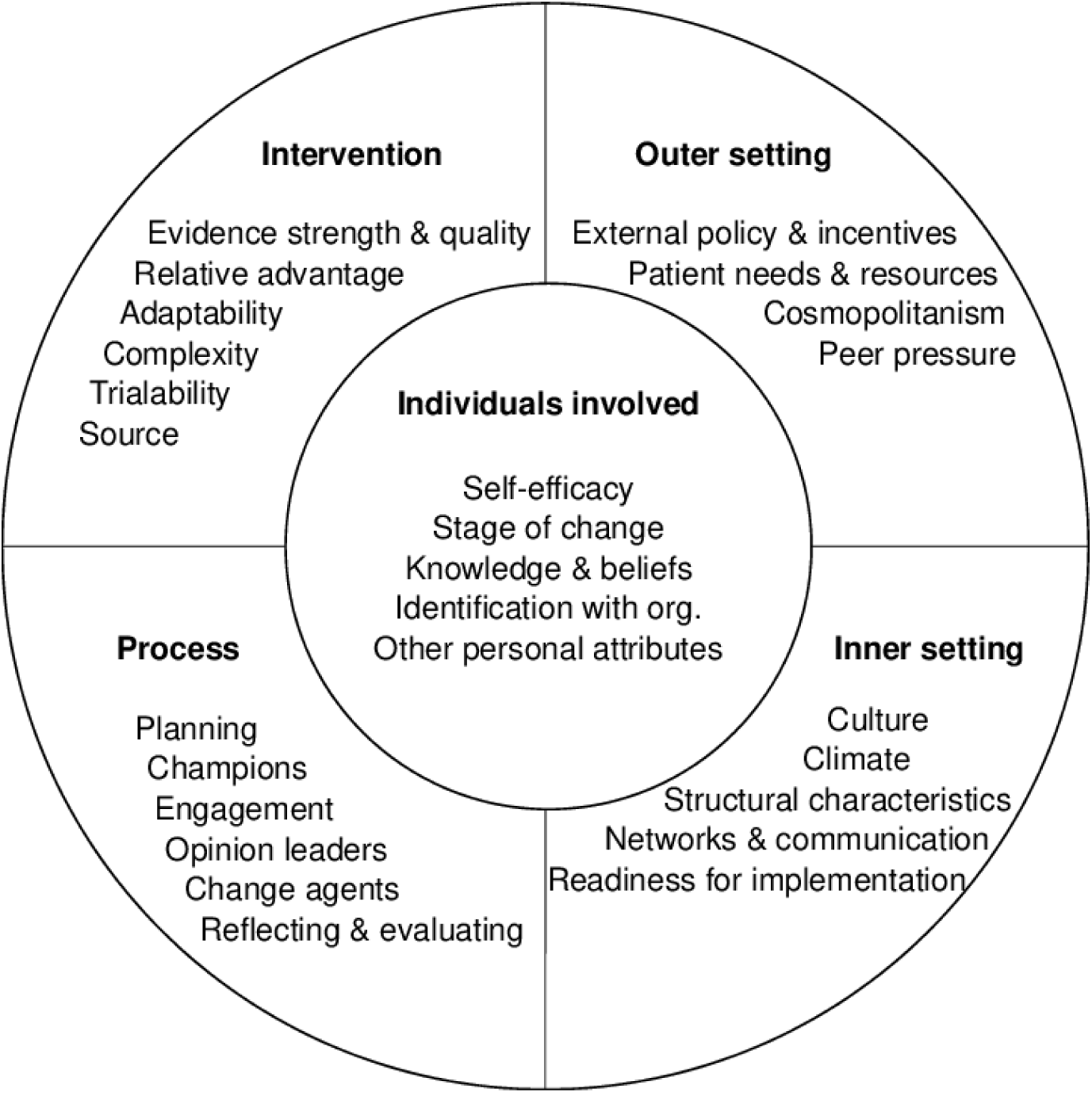
Consolidated Framework for Implementation Research (Cullen et al., 2022)

### Reflexivity

We considered the researchers’ backgrounds and experiences and how they influenced the design, conduct, data collection and analysis. VM had a background in psychology from India and had been in the UK for one year. English is not her first language, and she had no prior relationship with any of the interviewees at the time of the study. VM conducted the staff interviews bringing an outsider perspective to the UK dementia care context. PR and GL developed the DREAMS:START intervention and are experienced clinicians and researchers in dementia care in the UK NHS.

## Results

Ten interviews (30-75 minutes each) with healthcare professionals were conducted in June and July 2024 and nine with facilitators between February 2022 and May 2023, characteristics are presented in Table 1. Most participants were female (n = 16, 84%). The largest professional groups were clinical research assistants (n = 5, 26%), assistant psychologists (n = 4, 21%), and clinical psychologists (n = 4, 21%). Health care professionals were based mainly in older people’s mental health services (n = 7, 36%). The facilitators (n=9) delivered DREAMS:START to between 1 and 14 carers each, with a mean of 4.56 carers per facilitator (SD=3.71). Most of the healthcare professionals were White British (n=7).

**Table 1.** Participant characteristics.

| **Variable** | ***N=19 (%)*** |
| --- | --- |
| **Gender** |  |
| Female | 16 (84%) |
| Male | 3 (16%) |
| **Professional role** |  |
| Clinical research assistant | 5 (26%) |
| Assistant Psychologist | 4 (21%) |
| Clinical Psychologist | 4 (21%) |
| Mental Health Nurse | 3 (17%) |
| Service Manager | 1 (5%) |
| Occupational Therapist | 1 (5%) |
| Community mental health support worker | 1 (5%) |
| **Team setting** |  |
| Older peoples' mental health services | 7 (36%) |
| Clinical research department | 5 (26%) |
| Memory Service | 3 (16%) |
| Older peoples' community Mental Health Team | 2 (11%) |
| Older peoples' Integrated Care Team | 2 (11%) |

We identified five themes regarding common facilitators and barriers to implementation. These were: 1) perceived need for interventions targeting sleep and dementia, 2) importance of evidence-based practice, 3) service manager attitude and support, 4) interconnectedness with other professionals, and 5) attitudes toward change: staff and funding considerations.

### Theme 1 – Perceived need for interventions targeting sleep in dementia (CFIR – Patient Needs and Resources)

Professionals described how sleep disturbance in dementia is currently addressed and treated in services. They reflected on the limits of current support for carers and considered how DREAMS:START could address any unmet needs.

#### Sleep issues reported by patient-carer dyads

Participants highlighted how family carers were in urgent need of interventions to address sleep issues.

> *‘A lot of carers struggle with sleep. I had one lady, she got to the point where he (husband with dementia) just wasn’t sleeping, and she sat on the floor crying her eyes out one night and said I cannot cope anymore. If there’s something out there that can help, then, she’ll go with it’*. (Interview 7, F, MH Nurse)

Participants reported that many people with dementia had sleep impairment, which impacted them and their family, particularly nighttime waking. Other frequent issues included sleeping through the day, earlier bedtime, and getting dressed and moving around in the middle of the night.

> *‘It’s often waking in the night and thinking it’s daytime. Getting up, getting dressed, and then going downstairs… is one of the biggest issues.’* (Interview 2, F, Clinical Psychologist)

Disturbed sleep was considered to have a deleterious effect on cognitive and physical functioning of people with dementia and their family, who had interrupted sleep, anxiety and anticipatory worry due to their relative being up and potentially hurting themselves.

> *‘When you come to sleep, it’s vital to someone’s general health. But, if the person with dementia isn’t going to be getting any sleep it’s likely that they’re not going to be as cognitively well, even despite dementia’.* (Interview 4, M, Clinical Psychologist)

#### Current pathway for sleep problems

The primary function of NHS memory services was felt to be initial diagnostic assessments and reviews rather than intervention and treatment of symptoms like sleep disturbances.

> *‘Other things would end up being prioritized. So, I would imagine, for nurses, it would be things like initial assessments, and reviews. For support workers, it might be more about physical health monitoring.’* (Interview 4, M, Clinical Psychologist)

Following a dementia diagnosis, participants consistently observed sleep disturbances as a challenge. However, there was a gap in the post-diagnostic support available - only medication and some sleep hygiene advice was offered but no structured intervention.

> *So, I think at the point of diagnosis, if there are concerns about sleep, then the psychiatrist or the memory clinic nurse would give… I assume basic advice around sleep… it wouldn’t be an intervention or anything like an assessment. It would just be advice.’* (Interview 2, F, Clinical Psychologist)

Several participants discussed medication, specifically melatonin, zopiclone, and temazepam, which while often used were not judged to be a helpful solution.

> *‘Zopiclone… inadequate. You know, it might get them off to sleep, but they’re not staying asleep… the other one is Temazepam. Again, it is effective, but it leaves the patient with a hefty hangover the next morning and increases falls and anyway, we’ve got frail patients.’* (Interview 7, F, MH Nurse)

### Theme 2 – Importance of evidence based practice (CFIR -Intervention Characteristics, Evidence Strength & Quality, Relative Advantage and Cost)

Having a scientific evidence base was considered crucial to determine uptake, support and implementation of new interventions. Participants spoke about the importance of a strong evidence base from a clinical trial and of how cost-effectiveness and clarity of the referral process set competing interventions apart within the NHS.

> *‘I think it’s one of those, isn’t it, where you’ve got to prove that it works for them to be able to sign it off because they’re not going to invest that time if it ain’t going to work.’* (Interview 7, F, MH Nurse)

### Cost-effectiveness and return on investment

With funding required for the staff time to deliver DREAMS:START, two-day facilitator training, supervision, lightbox, and actigraphy watch, service managers would need clear data on cost-effectiveness and scalability.

> *‘The budget is always tight for us. I know that the lightbox that’s part of DREAMS:START… maybe, spending more money might be difficult. But I don’t think it would be impossible, it would be a case of justifying it.’* (Interview 2, F, Clinical Psychologist)

Different types of evidence perceived as necessary to win the support of clinical and operational staff. While the clinical staff needed evidence of efficacy, operational staff needed costing figures. Participants therefore suggested a presentation for operational staff, summarising the cost and benefit figures to secure their support.

> *‘If there is some sort of presentation, that we could then give to our service managers to be able to say-this is the cost, the potential benefit. I think that would help because we could propose that to service managers.’* (Interview 4, M, Clinical Psychologist)

#### Clear referral process

Participants suggested that DREAMS:START should only be used if carers were stressed or people with dementia’s sleep was significantly disturbed, to ensure that those who needed it most would receive it.

> *‘We would have to have some eligibility criteria in place so that the neediest patients and carers were prioritized… in terms of the carers, it’s experiencing the most significant carer stress, and for the people with dementia-those whose sleep was significantly disturbed.’* (Interview 6, F, Service Manager)

### Theme 3 – Service manager attitude and support (Inner Setting, Implementation Climate, Structural Characteristics)

Participants highlighted that service managers must weigh clinical evidence, cost considerations, and operational constraints to decide about intervention implementation.

#### Providing permissions and allocating budget

Service managers would have to decide to pay for training. To justify spending, they need to know about demand, including whether family carers are asking for such an intervention.

> *‘I would need to be able to persuade senior managers that it was going to be a really useful intervention. If we had clear evidence that there was a push from carers to receive this.’* (Interview 6, F, Service Manager)

However, interventions that need additional funding may lead to others being reduced and are considered carefully, ensuring there is a return on investment. Hence, budget holders might focus more on budgets than on needs expressed by service users.

> *‘I know that my service manager considers budget and staff capacity. We’re supposed to be patient-led, but I think sometimes the decisions that he makes are not always in the patient’s best interest, but about what he deems better for the service.’* (Interview 1, F, Assistant Psychologist)

#### Prioritising needs

Service managers in memory clinics prioritise meeting national targets, which currently focus on initial diagnosis within 6 weeks of a GP referral. This makes it more challenging to invest in post-diagnostic interventions like DREAMS:START.

> *‘Senior managers are focused on those diagnostic targets, which are the national targets. I think they value the fact that we are offering this post-diagnostic end-of-life service, but there are no national targets associated with that.’* (Interview 6, F, Service Manager)

Sometimes acute situations and crises, for example, aggression might be prioritised over people with sleep problems at home.

> ‘*I think it’s just, that there’s a lot of extra service pressures around working with behaviours that challenge and trying to provide interventions to those cases. And there’s an element of prioritization of clinical needs. Do we have a lot of cases where people are being physically aggressive in care homes, for example, that might then end up needing prioritization over people’s sleep difficulties in the home.’* (Interview 2, F, Clinical Psychologist)

### Theme 4 – Interconnectedness with other professionals (CFIR – Process, Engaging, Executing, Reflecting & Evaluating)

Participants spoke about collaborating with other professionals within their organization, to seamlessly integrate new interventions. They considered how the training, supervision, and peer support process would occur in practice and strategies to manage resource constraints.

#### Importance of training

As the intervention would be delivered by non-clinically trained psychology graduates, quality training procedures focused upon the clinical problem, the intervention and describing difficult clinical situations that may occur during delivery would be required to ensure structured, consistent, standardized delivery in the NHS which would cover the range of patient needs and presentations.

> *‘Make it [DREAMS:START training] intensive for us to be able to understand it, to be able to answer every question about how we deliver that out.’ (Interview 7, F, MH Nurse)*

> *‘So, it’s I think it’s a challenge to do this intervention where you have to try and get everyone involved. I think it would have been nicer during that [facilitator training] session to have pre-planned care and examples with more extremes… what do we say to someone who is reactive, more advanced? How would we adapt?’ (Facilitator 5, F, Clinical RA)*

Participants discussed how turnover rates of assistant psychologists, typically 1-2 years, would create additional training costs. A train-the-trainer approach might help wherein a group of experienced staff members would be trained in the intervention delivery and training procedures, who would then be responsible for cascading the training. This would enable trainers to customize the training to the specific service requirements while making training junior staff more economically viable:

> *‘You could have a train-the-trainer approach. You train people to train other people so they can then train their staff at no cost.’* (Interview 9, F, Clinical Psychologist)

#### Value of Supervision

Clinical supervision, necessary for facilitators of an intervention like DREAMS:START, was felt to be important but not necessarily a barrier to implementation. Supervision for facilitators is mandated by professional guidelines and already in place, so would not require additional resource.

> ‘*I don’t think that supervision would be a problem because they’re getting supervision anyway, so it would be included in that. And it wouldn’t take up extra time because… that time has already been set aside for supervision.’* (Interview 6, F, Service Manager)

The clinical judgment and guidance of supervisors was considered important for facilitators to focus the intervention and act appropriately when people with dementia and their carers would want to bring needs beyond sleep disturbances.

> ‘*I haven’t worked in the NHS before. So, all this is new to me, having my supervisor there to talk about these things… if a person’s going off in this way, what you should look out for, and this is how you can bring it back.’* (Interview 1, F, Assistant Psychologist)

The foregrounding of sleep issues in multi-disciplinary teams was considered beneficial. Hearing how other people tackle challenges would help participants feel confident in delivering the intervention themselves.

> ‘*When we did the dementia training previously, a lot of the shared information in MDTs about that in lots of meetings and one-to-ones was effective, was it not effective… We have a lot of open forums in terms of discussions that people can bring things to, and they’re not shy at bringing it to.’* (Interview 7, F, MH Nurse)

#### Team Champions

Champion individuals or teams, who drive change and innovation in the face of obstacles were considered important to initiate any change within the NHS, with a support system for the champions through support groups or troubleshooting sessions considered crucial.

> ‘*The challenge is having a member of staff who is supportive of DREAMS, who will be that champion. We see in our research teams when we have even one person, it makes such a difference in how a team engages with the intervention.’* (Facilitator 5, F, Clinical RA)

### Theme 5 – Attitudes toward change: Staff and funding considerations (CFIR – Compatibility, Relative Priority, Readiness for Implementation)

Considerations such as who delivers the intervention, staff capacity, job plans, funding in the NHS, and equipment cost and provision were discussed.

#### Who will deliver the intervention and where?

Participants agreed that support workers and assistant psychologists would be ideally placed to deliver this intervention. Capacity might be found as the NHS is a “moveable beast,” where dynamic factors impact staffing capacity. There could be gaps where the intervention would not be offered to people, due to assistant psychologists moving on and posts being empty.

> *‘Potentially, I think it’s always hard because services are incredibly stretched at the moment. Assistant psychologists are always moving on and we need to train new staff to keep up with the existing caseloads.’* (Interview 9, F, Clinical Psychologist)

Mode of delivery was considered in terms of quality of interaction and practicality. Most felt that in-person interventions, especially those conducted at the patient’s home where the facilitator could make recommendations based on the living environments worked the best. However, this was not always feasible and the intervention could be delivered online, with technology a challenge for some carers. Ultimately, participants judged that all methods of delivery have pros and cons and the decision should rest with the carer.

> *‘Delivering those sessions in patients’ or carers’ houses gives me a better point of view on what’s going on in the patient and the carer’s natural environment.’* (Facilitator 6, M, Support Worker)

> ‘*Sometimes doing things remotely was challenging, but mostly it was helpful for people because it meant that people were still able to receive the intervention. Carers didn’t want people to come to their homes, especially living with vulnerable people [during the Covid 19 pandemic].’* (Facilitator 4, F, Research Associate)

#### NHS funding considerations

Budget allocation was uncertain even for experienced clinical leads. “Extra” or non-essential spending was believed to be scrutinized, although it was not clear, how this was defined.

> ‘*Financially everything is restricted. So, I think finding any extra funding is tricky. I don’t think we’ll get too much extra funding.*’ (Interview 4, M, Clinical Psychologist)

Although initially wary about funding for the lightbox and actigraphy watch, participants judged that having a smooth equipment return process could make it possible.

> ‘*I don’t know how much these things cost, but I think if that was on a sort of loan basis, I think that would be OK if we could reuse those items.’ (Interview 6, F, Service Manager)*

#### Attitudes toward change

Although multiple factors must be considered, and NHS-wide implementation of an intervention is daunting, participants expressed optimism about the potential to implement DREAMS:START.

> ‘*The NHS is a galaxy unto its own, so these are just some of the things. So, people are very sober about the marathon. Do not give up on the marathon, it is worth the run. We just need to prepare.’* (Interview 3, F, Clinical Psychologist)

## Discussion

### Main Findings

In our pre-implementation qualitative study, we identified a high perceived need for DREAMS:START, due to sleep disturbances being common and distressing for people with dementia and their family carers, relative to a lack of effective and available interventions. Having a robust evidence base was important. Commissioners need cost-benefit analysis, cost-effectiveness data, and cost-saving (Marden et al., 2013) to determine whether an intervention can be implemented. DREAMS:START, based on a multicentre RCT is clinically and cost-effective, and cost-saving (Rapaport et al., 2024; Gonzales et al., 2025). We identified a need for a standardised presentation which explains aims, evidence and cost-effectiveness for DREAMS:START. This has now been designed, primarily for commissioners, service managers and clinical staff but to be accessible for families too. https://www.ucl.ac.uk/brain-sciences/psychiatry/our-research/mental-health-older-people/DREAMS:START.

Although the participants acknowledged the importance of cost-effectiveness, they were concerned that improving sleep was not perceived as a target in the NHS. A multidisciplinary approach to implementation was perceived as a key facilitator, via training, supervision and a ‘train-the-trainer’ approach, along with recognising local champions who would support and enhance implementation. Clinical champions have been regarded as central to implementing evidence-based practices in health settings (Byers et al.,2020). They would help in overcoming organizational barriers, and demonstrate practice change, leading others by example. Participants suggested a flexible and personalised model of delivery. Blended care is considered ‘the best of both worlds’ (Wentzel et al., 2016) as it enables personalization which could improve treatment adherence.

Participants discussed NHS’s capacity for change, expressing that while change is possible, it tends to be slow, thus reflecting broader systemic and organisational issues. Hierarchical structures, bureaucratic processes and prevailing organisational preference for sticking with existing protocols have been identified as barriers that can constrain staff behaviour and slow down procedural changes (Smith et al., 2024). Embedding new practices like DREAMS:START not only requires staff motivation, but also leadership-buy in. The COVID-19 pandemic showed how encouraging collaboration and reducing bureaucracy can accelerate decision making and implementation (Irons, 2024).

The value of diagnosing dementia is unclear if it is not followed by support for families to navigate its emotional, practical, and future-planning implications (Mansfield et al., 2022). In England and Wales, although the national Dementia Well Pathway (NHS England, n.d.) promotes support that is comprehensive and ongoing, service provision remains heavily concentrated around the point of diagnosis and the first year thereafter (Bamford et al., 2021). This limited focus creates a gap in sustained post-diagnostic support. Sleep disturbances are one of the primary reasons for people with dementia moving into 24-hour care (Gibson et al., 2023). This may be the only viable option for people with dementia and their families, but most people prefer to stay in their own homes if they can (Fleming et al., 2015). If families are given more support to deal with the psychological, behavioral, and emotional challenges of dementia, it might be possible to sustain care at home for longer (Edwards et al., 2018), thus easing the load on both the family unit and wider society.

People with dementia being able to stay in their own homes has been shown to generate greater cost savings, even if these benefits are not always realised at the service level. DREAMS: START saves money system-wide but increases local expenditure, which creates a perverse incentive for practices, discouraging adoption due to one-off equipment costs for lightboxes and actigraphy watches.

Overall, the themes identified in this study can be understood through the Consolidated Framework for Implementation Research (CFIR) domains - intervention characteristics, inner setting, outer setting and process. This highlights that organizational contexts and readiness are as important as intervention efficacy for implementing DREAMS:START.

### Implications

The study provides preliminary insights about facilitators and barriers to implementation. It will enable the implementation team to reflect on factors that make the uptake of an intervention as smooth as possible. The findings will be instrumental in the future scaling up of DREAMS:START across the NHS. This could fill a significant gap in post-diagnostic support, leading to enhanced quality of life for both people with dementia and their carers.

### Strengths and Limitations

This study has several strengths, including the use of the CFIR, a theoretically informed and structured approach to identify the key factors that influence the implementation of DREAMS:START. Feedback from stakeholders not involved in the initial trial alongside clinical practitioners who had delivered the intervention to family carers, ensures that the study offers a wider perspective on the practical challenges of real-world implementation. However, the study also has limitations. Ultimately, the people agreed to be interviewed were more interested in the intervention and saw merit to it, they were more likely to have an enthusiastic and optimistic point of view about implementation. Additionally, the study was conducted during a period of significant pressure on NHS services with increasing waiting lists and strikes, which may have influenced participants’ views on the feasibility of implementing new interventions. Service commissioners were approached for the study but did not agree to participate. As a result, our understanding of commissioning priorities and constraints had to be inferred from accounts of other NHS staff, such as service managers and clinical psychologists.

### Future Directions

Future research should focus on how to ensure smooth implementation of DREAMS:START, to use the intervention in a small number of NHS trusts, explore the impact of clinical champions to refine the delivery process and address practical challenges. The intervention could be adapted to be used with people speaking languages apart from English, to enhance inclusivity. It has already been translated into Hindi. Finally, ongoing evaluation of outcomes following the implementation of DREAMS:START will be essential to ensure that the intervention continues to meet the needs of those it is designed to support.

## Author Contributions

VM formulated the research questions, conducted participant recruitment, interviews, and led the data analysis. PR and GL supervised the study, contributed to the study design, interpretation of findings, and critical revision of the manuscript. All authors contributed to drafting or revising the article, approved the final manuscript, and agree to be accountable for all aspects of the work.

## Declaration of Interest

None

## Funding Statement

This work was supported by the National Institute for Health and Care Research (NIHR) Health Technology Assessment (HTA) Programme grant (NIHR HTA 128761). The views expressed are those of the author(s) and not necessarily those of the NIHR or the Department of Health and Social Care.

## Transparency Declaration

The lead author confirms that this manuscript is an honest, accurate, and transparent report of the study. No significant aspects have been omitted, and any discrepancies from the original study plan have been explained.

## Data Availability Statement

The data supporting the findings of this study are available from the corresponding author upon reasonable request. Due to confidentiality agreements, data are not publicly available.

## Notes

### Competing Interest Statement

The authors have declared no competing interest.

### Author Declarations

The study received ethical approval from the London-Camden & Kings Cross Research Ethics Committee (Reference: 20/LO/0894) in September 2020 with an amendment agreed in April 2024 to conduct additional interviews.

